# Physician Evaluations of Large Language Model-Generated Responses to Medical Questions by Region and Years in Practice: A preliminary study

**DOI:** 10.1101/2025.08.15.25333788

**Authors:** James Brooks, Paa-Kwesi Blankson, Peter Murphy Campbell, R Adams Cowley, Tsorng-Shyang Yang, Tijani Oseni, Anny Rodriguez, Muhammed Y. Idris

## Abstract

**Background:** Large language models (LLMs) have demonstrated a unique ability to generate clinically accurate responses to patient questions, in some cases outperforming physicians. However, little is known about how physician evaluations of such responses vary globally and by years in clinical practice.

**Objective:** This study builds on prior work by comparing LLM-generated and physician-authored responses to patient questions using two general-purpose LLMs in an international sample of physicians. Participants were asked to rank responses based on accuracy and responsiveness.

**Methods:** We conducted a survey to assess physician preferences for AI- and human-generated responses to patient questions from the r/AskDocs subreddit. Participants reviewed anonymized answers from ChatGPT-4.0, Meta.AI, and a verified physician, ranking each from best (1) to worst (3). We summarized respondent characteristics descriptively. The primary outcome was the mean rank of each response type. Sensitivity analyses included pairwise win proportions and full rank distribution visualizations.

**Results:** Fifty-two physicians completed the survey, most of whom were male (78.8%), aged 25–34 (53.8%), based in North America (48.1%) or Africa (25.0%), and over half (53.8%) had less than 5 years of clinical experience. Across all regions, ChatGPT-4.0 and Meta.AI responses were preferred over physician-authored responses, with ChatGPT-4.0 ranked highest in Africa, Asia, Asia Pacific, and North America, and Meta.AI slightly favored in Europe and the Americas. By years in practice, AI-generated responses consistently outperformed physician responses, with ChatGPT-4.0 most preferred among those with less than 15 years of experience and showing the greatest advantage in the 10– 15 year group.

**Conclusions:** In our global sample, most physicians preferred LLM-generated responses over those written by human contributors. However, preferences varied by geographic region and years in clinical practice, suggesting that both cultural and experiential factors shape physician attitudes toward Artificial Intelligence (AI). These preliminary findings highlight the need for larger, adequately powered studies to assess statistically significant differences and interactions across subgroups. Such research is essential to inform context-specific strategies for integrating AI into patient-facing communication.

**Trial Registration:** N/A

## Introduction

The ability of artificial intelligence (AI) and large language models (LLMs) to analyze large volumes of health data and generate clinically accurate, context-aware responses presents a transformational opportunity to improve healthcare.^1^ The adoption of LLMs in patient-facing clinical roles has progressed more slowly than in other areas, due in large part to concerns about bias and diagnostic errors in earlier models.^2^ Methodological advancements, such as instruction-tuning, retrieval augmented generation, and multi-step reasoning, have demonstrated significant improvements in both diagnostic accuracy and the clinical utility of LLMs.^3–5^ However, despite these advances, there remain mixed reactions in the medical community. Surveys reveal a mixture of optimism and caution among physicians regarding the integration of LLMs into clinical and research practice.^6,7^

Understanding how physicians perceive the accuracy, appropriateness, and usefulness of LLM outputs is essential not only for assessing clinical accuracy but also for building trust in these models. Many studies have conducted physician evaluations of LLM-generated responses to patient questions, which play a crucial role in shaping broader perceptions and offer insight into both clinical utility and professional acceptance. Some have found that physicians rated LLM-generated responses more favorably than those written by humans.^8,9^ While others have found that physicians rated LLM-generated responses more favorably than those written by humans, in specialized therapeutic areas like rheumatology.^10^

These mixed results underscore the need for more refined evaluations of LLM performance in medical question and answer tasks. In this study, we explore how physician evaluations of such responses vary globally by geographic region and by years in clinical practice. Specifically, we surveyed an international sample of physicians to assess anonymized responses from verified physicians and two general-purpose LLMs based on accuracy and responsiveness. In this short paper, we report mean rankings of each response type and explore variation by clinician background. In doing so, we demonstrate how contextual factors and professional experience shape evaluations of LLM-generated content, highlighting the need for larger, adequately powered studies to rigorously assess these differences.

## Methods

### Study Design and Participant Recruitment

We conducted a cross-sectional survey between March to May 2025. Eligible participants were licensed physicians and equivalent medical professionals and were recruited through institutional contacts, professional networks, and physician email distribution lists to ensure global representation. Participants were asked to review and ranked order anonymized answers generated by two large language models (ChatGPT-4.0 and Meta.AI) and by verified human physicians.

Medical questions and physician-authored responses were sourced from Reddit’s r/AskDocs forum, one of the largest public repositories of health-related queries, with over 689,000 members as of April 2025.^11^ This subreddit features responses from verified medical professionals, who are credentialed through moderator verification (e.g., submission of medical ID or diploma). Eighty-six questions and answers (from 300 of the most recent and highly rated physician responses) were selected based on clarity, generalizability, and the absence of identifiable information. From this subset, 30 question– answer pairs were randomly selected using numbergenerator.org. All data collection procedures adhered to Reddit’s terms of service.

Finally, for each of the 30 selected medical question–answer pairs, new responses were generated using ChatGPT-4.0 and Meta.AI in isolated sessions to avoid contextual contamination. As such, each question had three responses: one from a verified physician, one from ChatGPT-4.0, and one from Meta.AI. Participants were asked to rank each set of three responses to a medical question based on overall preference (1 = most preferred, 3 = least preferred). Respondents were asked to rank responses based on their perceived accuracy and responsiveness.

### Survey Content and Administration

The survey was constructed and administered using Qualtrics. Participants were asked to provide information about their primary specialty of practice, years in clinical practice, current level of practice, age, gender, and primary geographical region of practice. Participants were also asked to indicate their current reliance on AI in routine clinical practice. Each respondent was then presented with three anonymized responses (ChatGPT-4.0, Meta.AI, or verified physicians) to three randomly selected medical questions from a bank of 30. The order of the responses was randomized to reduce potential ordering bias, and all sources were anonymized to prevent identification.

### Statistical Analysis

Our primary outcome was the mean rank assigned to each system (1 = most preferred, 3 = least preferred) across all evaluation tasks. To examine subgroup variation, we compared the mean rank for each response type (ChatGPT, Meta.AI, verified physician) across categories of years in clinical practice and geographic region. To assess robustness, we conducted two sensitivity analyses: (1) the proportion of wins in pairwise comparisons between systems, and (2) full rank distribution visualizations. Given limited sample sizes within subgroups, no formal statistical comparisons were performed. All statistical analyses were conducted using R using version 4.1.2.

### Ethical Considerations

This study was reviewed by the Institutional Review Board of the Morehouse School of Medicine and determined to be exempt from full review due to its minimal risk (MSM-IRB 2276674-1). Participation was entirely voluntary, and all responses were collected anonymously. No personally identifiable or protected health information was gathered at any stage of the study.

## Results

A total of 52 physicians completed the survey, with nearly half (48.1%) based in North America and the rest distributed primarily across Africa (25%) and Asia (15.4%). Most respondents were young, with over half (53.8%) aged 25–34, and most (53.8%) had less than five years of clinical experience. The sample was predominantly male (78.8%). In terms of specialties, Surgery was the most common (26.9%), followed by Family Medicine and Anesthesiology (each 17.3%). Other specialties were less represented. On average, physicians reported integrating artificial intelligence into approximately 19.1% of their routine clinical practice (SD = 19.6), indicating varied but limited current adoption across the sample.

Table 2 displays the mean rank assigned to each response type across all physician evaluations, with lower scores indicating greater preference. On average, responses generated by ChatGPT-4.0 received the most favorable ratings, with a mean rank of 1.63 (SD = 0.68, 95% CI: 1.52–1.74). Meta.AI responses followed with a mean rank of 1.83 (SD = 0.72, 95% CI: 1.71–1.94), while verified physician-authored responses were ranked least favorably, with a mean of 2.53 (SD = 0.76, 95% CI: 2.40–2.65). These results suggest a clear overall preference among surveyed physicians for LLM-generated responses over those written by verified physicians.

**Table 1.** Demographic and professional characteristics of the physician survey respondents (n = 52)

| Variable | Number (%) |
| --- | --- |
| Age Category |  |
| 25-34 years | 28 (53.8) |
| 35-44 years | 14 (26.9) |
| 45-54 years | 5 ( 9.6) |
| 55-64 years | 4 ( 7.7) |
| 65+ years | 1 ( 1.9) |
| Sex |  |
| Male | 41 (78.8) |
| Female | 11 (21.2) |
| Specialty |  |
| General | 2 ( 3.8) |
| Surgery | 14 (26.9) |
| Family Medicine | 9 (17.3) |
| Internal Medicine | 5 ( 9.6) |
| Anesthesiology | 9 (17.3) |
| Emergency Medicine | 3 ( 5.8) |
| Pediatrics | 2 ( 3.8) |
| Psychiatry | 3 ( 5.8) |
| Other | 5 ( 9.6) |
| Years in practice |  |
| 10-15 years | 6 (11.5) |
| 5-10 years | 9 (17.3) |
| Less than 5 years | 28 (53.8) |
| More than 15 years | 9 (17.3) |
| Geography |  |
| Africa | 13 (25.0) |
| Americas | 1 ( 1.9) |
| Asia | 8 (15.4) |
| Asia Pacific | 3 ( 5.8) |
| Europe | 1 ( 1.9) |
| North America | 25 (48.1) |
| Other | 1 ( 1.9) |

**Table 2.** Mean Rank of Each Response Type.

| Mean Rank of Each Response Type |  |  |  |  |  |  |
| --- | --- | --- | --- | --- | --- | --- |
| Lower scores indicate greater preference |  |  |  |  |  |  |
| Response Type | n | Mean Rank | SD | SE | 95% CI (Low) | 95% CI (High) |
| ChatGPT | 150.00 | 1.63 | 0.68 | 0.06 | 1.52 | 1.74 |
| MetaAI | 150.00 | 1.83 | 0.72 | 0.06 | 1.71 | 1.94 |
| Physician | 150.00 | 2.53 | 0.76 | 0.06 | 2.40 | 2.65 |

Figure 1 presents the mean ranking of each response type, stratified by geographic region, along with associated error bars. Across all six regions, ChatGPT-4.0 and Meta.AI were consistently ranked more favorably than physician responses. In Africa, Asia, Asia Pacific, and North America, ChatGPT-4.0 held the lowest mean rank, indicating the strongest overall preference. In the Americas and Europe, Meta.AI slightly outperformed ChatGPT, though both LLMs were clearly preferred over human-authored responses. Physician responses received the highest (least preferred) rankings in every region, with particularly low ratings in the Americas and Europe. While the overall pattern of preference for LLM-generated content was consistent, small regional differences in the relative performance of ChatGPT-4.0 and Meta.AI suggest some variation in how physicians across different regions evaluate AI-generated responses.

**Figure 1.**
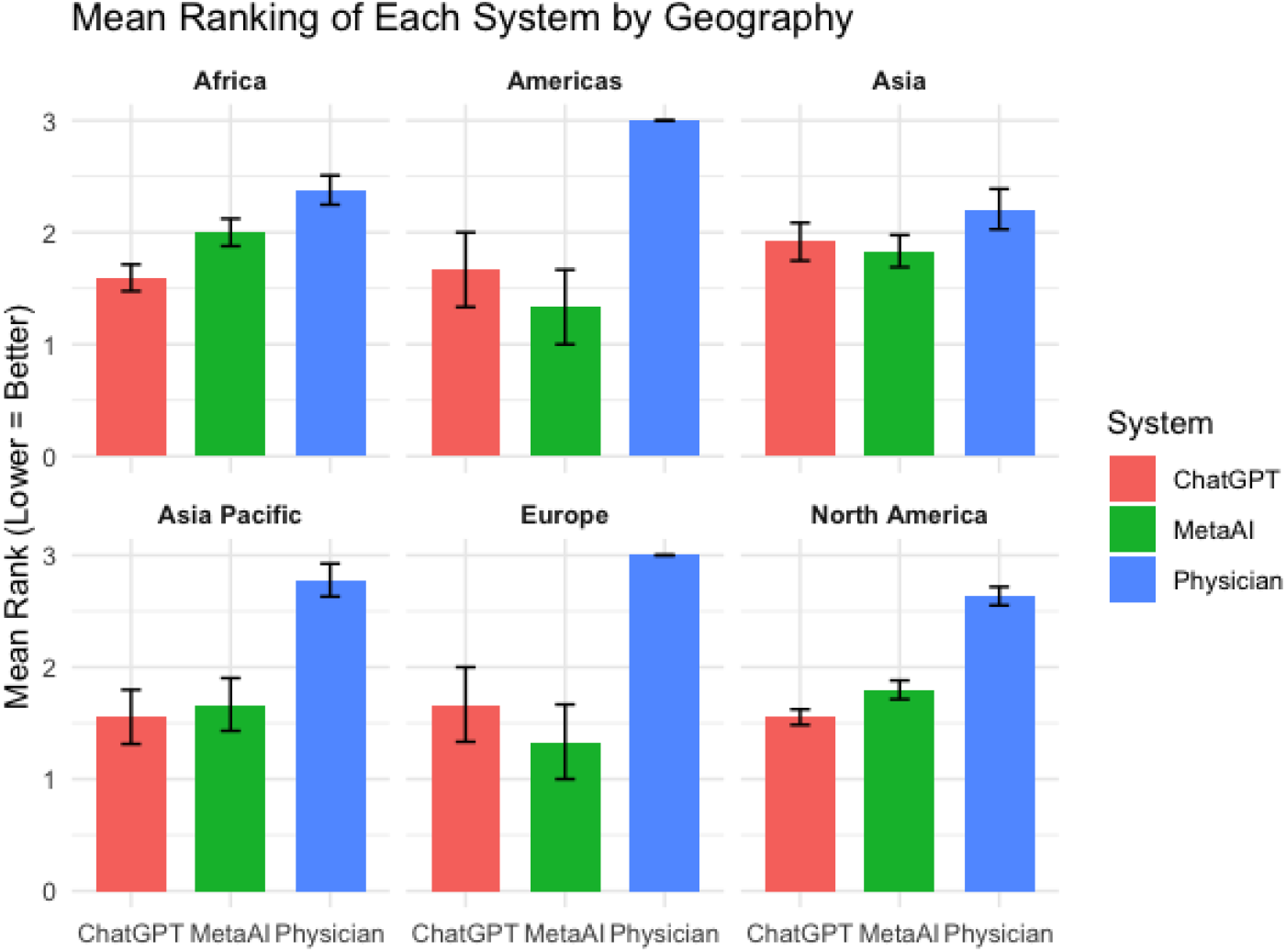
Mean Ranking of Each System by Geography.

Figure 2 displays the mean ranking of each response type stratified by years in clinical practice, again with lower values indicating greater preference. Across all experience levels, physician responses consistently received the least favorable rankings. Among participants with less than 5 years of experience, ChatGPT-4.0 was ranked most favorably, followed closely by Meta.AI. This trend was consistent across other experience categories, including 5–10 years, 10–15 years, and more than 15 years, where both LLM-generated responses were preferred over those written by verified physicians. Notably, the relative advantage of ChatGPT-4.0 over Meta.AI was most pronounced among those with 10–15 years of experience.

**Figure 2.**
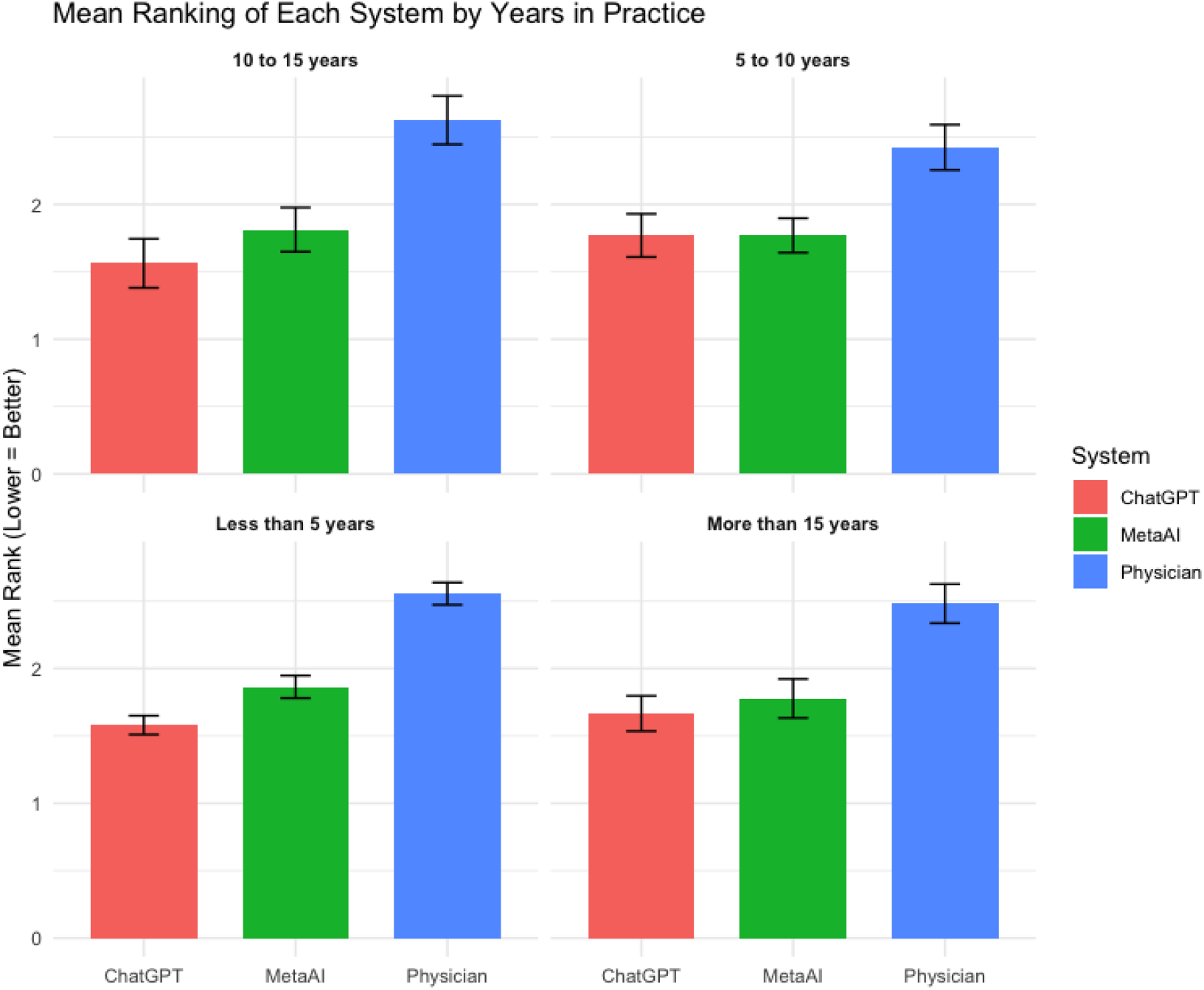
Mean Ranking of Each System by Years of Practice.

Results were consistent across sensitivity analyses. Pairwise comparisons confirmed the relative ordering of systems, and rank distributions revealed moderate variability across subgroups (see Supplementary Figures 1–2).

## Discussion

### Principal Results

In this exploratory study we demonstrate that LLM-generated responses were consistently preferred over those written by verified human physicians. Overall, ChatGPT-4.0 received the most favorable mean rankings, followed by Meta.AI, with physician-authored answers ranking lowest. This pattern held across geographic regions and years in clinical practice, with particularly strong preferences for ChatGPT-4.0 in Africa, Asia Pacific, and North America, and for Meta.AI in the Americas and Europe. Similarly, regardless of clinical experience level, both LLMs outperformed physician responses, with ChatGPT-4.0 maintaining a consistent advantage. Sensitivity analyses confirmed the robustness of these findings, supporting the overall conclusion that physicians generally favored LLM-generated medical responses.

### Limitations

This study has several limitations that should be considered when interpreting the findings. First, the relatively small sample size limited statistical power, especially for subgroup comparisons by geography and years in practice. As a result, observed differences in preferences should be interpreted as descriptive rather than inferential. This is especially true for physician responses from Europe and the Americas, which each only had 1 respondent. Second, the convenience sampling approach and voluntary participation may introduce selection bias, as physicians with stronger opinions or greater familiarity with (and interest in) AI tools may have been more likely to respond. Third, while the use of real-world patient questions from r/AskDocs enhances ecological validity, the specific platform and format may not fully reflect the complexity of clinical communication in real-world practice, nor are physician responses to questions on social media. Future studies with larger, more diverse samples and mixed-methods approaches are needed to validate and expand upon these preliminary findings.

### Comparison with Prior Work

The findings of this study both confirm and extend prior research around physician evaluations of LLM-generated versus physician responses to medical questions. Consistent with recent work we find that physicians generally preferred responses produced by models such as ChatGPT-4.0 and Meta.AI over those authored by clinicians.^8,9^ This trend was observed across all examined subgroups. Not all prior studies, however, found a clear preference for LLM-generated responses. Studies have shown that ChatGPT’s medical answers can be incomplete, inaccurate, or misleading when compared to clinician responses.^10,12,13^ Others have found a significant performance gap (18% drop in correctness, 29% lower consistency, and a 13% decrease in verifiability) in non-English medical question answering, highlighting substantial limitations in the reliability and quality of LLM-generated health information across languages.^14^ The mixed results highlight the context-dependent and variable nature of LLM performance and physician evaluations.

## Conclusions

Though exploratory, our study contributes valuable evidence to the expanding literature indicating that LLM-generated medical answers can be considered clinically acceptable by physicians. This suggests potential for LLMs to assist in patient education and clinical decision support under appropriate conditions. Our findings also underscore that physician perceptions of LLM-generated responses are not uniform but rather nuanced and influenced by various demographic factors (e.g., geographic region) as well as professional characteristics (e.g., years of clinical experience). These moderating factors may shape how clinicians evaluate the comprehensiveness, accuracy, readability, and overall usefulness of AI outputs. Such variability highlights the importance of tailoring AI integration strategies to specific user groups and clinical contexts, rather than assuming broad or universal acceptance. Therefore, while supporting the growing enthusiasm about the promise of LLMs, our study also calls for continued, rigorous evaluation of AI systems across subgroups and by extension settings to ensure safe, effective, and responsible adoption.

## Data Availability

All data produced in the present study are available upon reasonable request to the authors

## Acknowledgements

JB contributed to the conceptualization and methodology of the study, managed data curation, and participated in writing – review and editing. PKB contributed to data curation and writing – review and editing. AC, TSY, and TJ were involved in data curation. AR contributed to the conceptualization of the study. MYI played a leading role in conceptualization and methodology, contributed to writing – review and editing, and secured funding for the project.

## Conflicts of Interest

None declared

## Abbreviations

AI: Artificial Intelligence
JMIR: Journal of Medical Internet Research
LLM: Large language model
RCT: randomized controlled trial

## Multimedia Appendix 1

Multimedia appendices are supplementary files, such as a PowerPoint presentation of a conference talk about the study, additional screenshots of a website, mpeg/Quicktime video/audio files, Excel/Access/SAS/SPSS files containing original data (very long tables), and questionnaires. See https://jmir.zendesk.com/hc/en-us/articles/115003396688 for further information. Do not include copyrighted material unless you obtained written permission from the copyright holder, which should be uploaded together with your Publication Agreement form as supplementary file.

## Notes

### Competing Interest Statement

The authors have declared no competing interest.

### Funding Statement

This study did not receive any funding

### Author Declarations

Morehouse School of Medicine Institutional Review Board

